# Spatiotemporal Forecasting of Opioid-related Fatal Overdoses: Towards Best Practices for Modeling and Evaluation

**DOI:** 10.1101/2024.01.03.24300803

**Authors:** Kyle Heuton, Jyontika Kapoor, Shikhar Shrestha, Thomas J. Stopka, Michael C. Hughes

## Abstract

To inform public health interventions, researchers have developed models to forecast opioid-related overdose mortality. However, these efforts often have limited overlap in the models and datasets employed, presenting challenges to assessing progress in this field. Furthermore, common error-based performance metrics, such as root mean squared error, are not directly suitable to assess a key modeling purpose: the identification of priority areas for public health interventions. We recommend a new intervention-aware performance metric and establish a set of baseline models with competitive performance. To show how model and metric choice vary across locations, we explore two distinct geographies: Cook County, Illinois and the state of Massachusetts. We introduce a new, intervention-aware evaluation metric: the Percentage of Best Possible Reach (%BPR). The top-performing models based on error-based metrics recommend fixed-budget interventions in areas that do not always reach the most possible overdose events. In Massachusetts the top models, as ranked by our proposed %BPR metric, could have reached 18 additional fatal overdoses per year in our 2020-2021 test period compared to models favored by error-based metrics, assuming the ability to intervene in 100 census tracts out of the 1620 in Massachusetts. We release open code and data for others to build upon.

Repository for code and data: https://github.com/tufts-ml/opioid-overdose-models

## Introduction

The opioid overdose epidemic in the United States has resulted in over 450,000 deaths during the past eight years, with more than 80,000 fatal opioid-related overdoses during 2022, the highest yet in a single year.^1^ Managing the opioid overdose epidemic requires a constellation of efforts ranging from substance use treatment programs offering medications for opioid use disorder (e.g., methadone, buprenorphine),^2,3^ harm reduction programs with provisions for overdose education and naloxone distribution, and comprehensive mental health and social support services.^4–6^ Beyond the provision of harm reduction and healthcare services, it is critical for policymaking to address the ever-evolving substance use environment and plan for targeted interventions.

There has been considerable variation in the availability of different types of opioids and the consequent increase in opioid use disorder and opioid-related fatal overdoses in the past two decades. The current fatal opioid overdose epidemic has been characterized by four waves.^7,8^ In the early 2000s, prescription opioids drove overdoses. Then, heroin-related deaths surged post-2010, followed by a fentanyl spike in 2013.^9^ This culminated with the fourth wave of combined stimulant and fentanyl-related overdoses.^8^ These shifts in supply accompanied changes in social and ecological conditions, impacting substance use behaviors in varying ways across geographic regions.^10,11^ Hence, it is critical to examine local spatiotemporal variation in opioid overdose outcomes, identifying the most-impacted areas and predicting future outcomes to inform preemptive public health responses.

A growing body of research^12^ has explored spatiotemporal variations in the opioid overdose landscape. Yet *forecasting* approaches are in a nascent stage and there are few prediction studies at the population level^13^. Other research focuses on patient-specific risk prediction ^14–16^, assuming access to detailed, person-level demographic and medical history data. However, there are immense challenges in compiling rich datasets for person-level analysis. Most state-level public health authorities may not have access to the data and technical resources to conduct individual predictive modeling. Analyses focused on population-level predictions that solely depend on more readily-available aggregated data have the potential to be more easily adopted by public health authorities with limited resources.

While a number of prior studies have identified historical overdose “hotspots”^17–19^, fewer studies have forecasted future spatiotemporal overdose spikes. Research that focuses on hotspots often assumes that identified clusters represent the locations where the highest needs will exist in the future. In our analyses, we show that this assumption does not always hold. Intervention and policies that rely on such findings may be acting on lagged measures of opioid burden, thereby limiting the effectiveness of interventions. Existing research also spans a broad range of spatial and temporal resolutions. In geographic space, studies range from coarser county-level analysis^17^, to finer analyses based on ZIP Codes, census tracts, or census block groups^20^. Temporally, studies range in focus across yearly aggregated^17^ data, quarterly^21^, or weekly data^22^.

The overarching goal of our study is to help public health departments make short-term forecasts of future overdose events to enable planning of geographically and temporally targeted interventions that are cognizant of limited resources and needed intervention efficiencies. We focus specifically on development of forecasts at a fine spatial scale (census tracts) at annual intervals, which we selected to match the decision-making needs of public health agencies. In order to understand the role of different forecasting models and evaluation metrics on different communities, our evaluations cover two distinct catchment areas. First, we study Cook County, Illinois, covering over 5 million residents of Chicago and its surroundings, where we forecast death events across 1328 populated census tracts from years 2015-2022 through analysis of publicly available data. Second, we study the state of Massachusetts, where we forecast fatal overdose events across 1620 census tracts representing over 6 million residents from 2001-2021. These locations were selected based on data availability, and to demonstrate the impact of model and metric choice at multiple locations.

To establish best practices for modeling and evaluation, we carefully compare different modeling approaches and performance metrics in each catchment area. We implement a comprehensive set of existing models – including heuristic baselines, statistical models,^12,20,22,23^ and neural networks^22^. We then assess the opioid-related fatal overdose forecasts they produce for both Cook County and Massachusetts at the census-tract-level at annual timeframes. We compute widely-used error-based performance metrics and introduce a new intervention-focused performance metric. Our Python-based software is available for other researchers to reuse and build upon: https://github.com/tufts-ml/opioid-overdose-models.

## Methods

### Data Sources and Preparation

To assess models, we assembled two datasets suitable for forecasting opioid-related fatal overdoses annually at the census tract level. Our relatively coarse annual temporal scale was chosen to match the frequency at which decision-makers might set new priorities and at which new reliable data become available. We chose the census tract spatial scale due to its potential for targeted interventions at a sub-municipality level. Each census tract by design contains a mean count of 4000 people (with a range of 1200-8000^24^). For many (but not all) interventions, costs scale with population size, and thus the cost of deploying an intervention in any tract is roughly uniform.

### Data source 1: Cook County, Illinois

We obtained fully de-identified data from the Cook County Government Medical Examiner Case Archive^25^ for opioid-involved overdose deaths from August 2014 (the first date records are available) to May 2023. These data contained every fatal incident under the medical examiner’s jurisdiction that was determined to have any opioid as a primary cause. We used the provided incident latitude and longitude to map each overdose fatality to one of 1328 census tracts. Because the underlying data is in the public domain, we make our processed Cook County data available in our shared repository.

### Data source 2: Massachusetts

We obtained death certificate data from the Massachusetts Registry of Vital Records and Statistics for opioid-involved overdose deaths between 2001 and 2019. These deaths were defined as unintentional, intentional, and undetermined drug poisonings containing an opioid code (ICD-10 codes T.40.0-T40.4, or T40.6) as a “multiple cause of death”. Each fatal overdose is linked to a calendar date as well as a residential street address. Decedent addresses for the place of residence at the time of the fatal overdose were geocoded, assigning latitude and longitude measures to each event that was then mapped to one of 1620 census tracts using the 2020 census tract boundaries.

### Dataset Preparation

For each dataset, we computed the observed number of fatal overdose events *y_s,t_* at time unit *t* for individuals residing in spatial tract *s*. We employed open tools^26^ that utilize the US Census Geocoding API to map locations (street address or latitude/longitude) to its corresponding census tract, using the tract boundaries for the states of Massachusetts and Illinois defined by the U.S. Census Bureau in 2020.

In each dataset, a uniform set of covariates is available as input for prediction models. At each time period *t* and spatial region *s*, we provide the history of fatal overdose counts from previous times in that region, as well as the spatial location (numerical latitude and longitude of the tract), and timestamp (numerical time, measured in years since the first available year for that dataset). Further, for each census tract *s* at time *t*, an optional covariate vector represents social vulnerability across socioeconomic status, age-related demographics, minority status, housing, and composite dimensions using percentile ranking across all census tracts in that state. These features stem from the five dimensions of the Social Vulnerability Index (SVI)^27^ published between 2000 and 2018 for every U.S. state. Published tract values are updated every five years; we selected the closest value to each time period t. These SVI features were chosen for their simplicity and portability, mirroring the role of higher-dimensional socioeconomic covariates in previous studies ^23,28^.

## Metrics

To evaluate model forecasting accuracy against observed mortality in a test period, suitable performance metrics were essential. Our study considered both commonly-used error-based metrics and a new intervention-focused metric.

### Error-based metrics

Model performance is often assessed via summary statistics of the errors between predicted and observed mortality across all S spatial regions in the test period. Within this category, two common metrics are Root Mean Squared Error (RMSE) and Mean Absolute Error (MAE), both defined in Equation 1 below. RMSE calculates the square root of the average squared errors, while MAE computes the average of absolute errors.

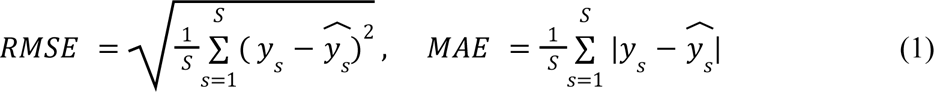

Both RMSE and MAE have been concretely used as the primary metrics to assess opioid overdose forecasting^22,23,29^. RMSE can be more sensitive to large errors due to its use of squaring.

### Intervention-focused metric

In our intended use case of mitigating the opioid crisis, stakeholders at a public health agency could use a forecasting model to select a targeted subset of all possible census tracts in which to deploy an intervention in the near future. We assume these actors have a limited budget, allowing intervention deployment in a maximum of K of the S regions in their jurisdiction. For a given model, we can obtain its recommended set of K regions, which we refer to as the intervention set *I*, in two steps. Step 1: predict mortality counts for all S regions in the test period. Step 2: identify the K regions with the K highest predictions (breaking ties at random), and store these as the recommended set *I*.

To evaluate such intervention recommendations, we need new metrics. The error-based metrics defined earlier (RMSE or MAE) aggregate error for all S regions equally. They do not directly measure if a forecast’s guess of the K highest-risk areas specifically aligns with the actual areas with highest mortality. We thus wish to design a metric better aligned with how stakeholders will determine and assess intervention priorities. We suggest a model is favorable if, during the test period, the total count of fatal overdose events in its recommended set *I* of K tracts is as large as possible. This would indicate the model is good at identifying where adverse events will occur, and thus increase the possibility that stakeholders could “reach” and hopefully mitigate these events via interventions targeted at the model’s recommended tracts.

Our proposed metric, “best possible reach” (BPR), assesses a model’s recommendations via a ratio of two numbers. First, the numerator counts how many opioid-related fatalities actually occurred in the model’s recommended set *I* of size K. Second, the denominator counts the total number of opioid-related fatalities in the K regions that would be chosen with perfect hindsight of the actual count vector y = [y_1_, …, y_S_] of fatalities in the test period. Mathematically, we define BPR as follows

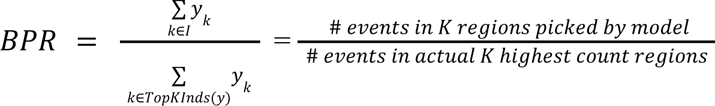

where TopKInds(y) denotes a function that returns the distinct indices of the K largest elements of vector *y*.

For public health applications, BPR holds a practical interpretation as the proportion of fatal overdose events the current model’s interventions would reach, compared to the perfect foresight of future events. BPR’s numerical value by definition will have a minimum of 0.0 and a maximum of 1.0. We typically convert the fractional BPR to a percentage ranging from 0-100%, denoted as %BPR. A higher %BPR value signifies a more effective model at deciding where to intervene. A value of 100% indicates perfect decision-making given a limited budget: there is no other set of K regions any model could have recommended that would reach more events.

Although independently developed by our team (see our preliminary workshop paper^21^), our proposed BPR metric closely resembles the metric suggested by a recent pre-registered trial^30^ and a later feasibility study^20^ to evaluate opioid overdose forecasts in Rhode Island. The primary distinction lies in the denominator: our BPR sums only the top K indices, while the alternative includes all S regions. We prefer our definition due to %BPR’s consistent range of 0-100%, with 100% representing a model that could not have made a better decision given the limited budget of K regions. In contrast, the alternative definition’s maximum value fluctuates based on observed data in the test period, making it difficult to know if another model could have done better.

## Models

Below we define a range of possible forecasting methods that can be used for our where-to-intervene prediction task. All methods are trained and evaluated via a common protocol using the same provided splits (train/validation/test) of the two available datasets (Cook County and Massachusetts). Thorough details about model fitting and hyperparameter tuning are provided in the Supplement. This study was reviewed by the Tufts University Health Sciences Institutional Review Board and deemed to be non-human subjects research.

### Simple Baseline Models

We study several easy-to-implement baseline models to highlight their comparative strengths. Public health practitioners seeking data-driven allocation of scarce intervention resources without sophisticated modeling could easily use these approaches.

Our first baseline, dubbed *all zeroes,* predicts uniformly across all S tracts that zero fatal overdoses will occur in the test period. This model, by definition, ranks all tracts as equally high-risk, so for metrics like BPR that require a set of K recommended regions we report an average over many samples of K distinct regions selected uniformly at random.

Our second baseline, known as *last year,* predicts that the mortality at the next year in a specific location will mirror the mortality observed at the most recent recorded year for that location.

The final baseline we consider is a *historical average,* which predicts the next timestep’s mortality as an average of all mortality counts observed over the preceding W timesteps. Here, the appropriate “look back” period length W serves as a model hyperparameter.

### Complex Models

Next, we consider several more flexible models with parameters that can be fit to the data. The first is the *weighted historical* average: a weighted average of the previous W years of overdose events. This is more flexible than *historical average*, because each year’s count is multiplied by a customized weight coefficient.

The next model we consider is a Generalized Linear Model (GLM) with a Poisson likelihood. This model assumes that fatal overdose count *y* for spatial tract *s* at time *t* is modeled by a Poisson distribution where the log of the mean parameter is a linear function of the covariate vector *x* for that tract and time:

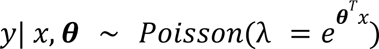

GLMs have limited flexibility due to the assumption of a monotonic relationship between covariates and fatal event count. We thus also include a Gradient Boosted Trees^31^ model, a popular more flexible ensemble of regression trees. Previous studies of opioid forecasting ^20,32^ have used similar tree ensembles.

In addition, we include three spatially-sophisticated statistical models used in recent opioid overdose forecasting applications. First, we include a *Gaussian Process* model^20^ for its ability to flexibly capture spatial and temporal correlations. We use similar covariance functions (“kernels”) to prior overdose forecasting work (details in the supplement). Next, *Bayesian Spatio-Temporal (BST)* models^23^ use a Markov Random Field to model inherent spatial and temporal trends. Thirdly, *NBSpLag* denotes a negative binomial regression model with spatially lagged features^28^, where each tract is informed by its spatial neighbors. In a variable selection experiment^28^, these spatially lagged covariates were found to be the most predictive features. Unlike previous evaluations of both *BST*^23^ and *NBSpLag* models, our study compares to the rich set of baselines described above.

Finally, we include *CASTNet*^22^, a neural network approach custom developed for opioid-overdose forecasting. Unlike previous methods, CASTNet employs multi-head attentional networks that allow predictions at a given location to be informed by learned “communities” of regions.

### Experimental Protocol

We applied each of the models described above separately to the Cook County, IL dataset and the MA dataset. In each case, we sought to use available historical counts of opioid-related fatal overdoses (together with other covariates described above) to predict future fatal overdose counts in each census tract. We further assessed how these predictions can be used to recommend where to intervene in the near future.

For training on a dataset, for all S regions we assemble covariate vector, fatality count pairs (*x_s,t_*, *y_s,t_*) for each year in the training set (t = 2010-2018 for Massachusetts, 2015-2019 for Cook County). The historical covariates inside each *x* vector can summarize the recent history of *W* previous years (W=10 for Massachusetts, W=5 for Cook County). Hyperparameters are chosen to maximize performance as assessed by BPR on a validation set of data from the year prior to evaluation (2019 for Massachusetts, 2020 for Cook County. Finally, models are evaluated on predictions for the final two years (2020-2021 in Massachusetts, 2021-2022 in Cook County).

From each model, we obtained predictions for each of the S tracts in each test year. We then computed each evaluation metric (RMSE, MAE, BPR) as well as an interval that quantifies our uncertainty in its precise value. Inspired by resampling methods for uncertainty quantification^33^, for each test year we obtained 50 different without-replacement samples of 1370 of the 1620 locations in MA (1078 of the 1328 in Cook County, IL), and recorded all metrics of interest for each sample. Our reported intervals quantify the min-max range of these 50 samples. We chose the number of tracts retained in each catchment area so that each sample on average retains 85% of all fatal overdose events.

## Results

Results from the experiments conducted on Massachusetts and Cook County data are summarized in Table 1 and Table 2, respectively. The best model(s) can vary depending on the chosen evaluation metric.

**Table 1.** Comparison of fatal opioid-related overdose prediction models trained on Massachusetts decedent data from 2010-2019, then evaluated on data from 2020 and 2021.

| Model | Metric |  |  |  |
| --- | --- | --- | --- | --- |
|  | Total overdoses identified in top 100 tracts | %BPR (K=100) | MAE | RMSE |
| All Zeros | 124.1 | 25.1, (24.8-25.8) | 1.24, (1.18-1.30) | 1.92, (1.83-2.01) |
| Last year | 254.5 | 51.8, (49.5-54.4) | 1.09, (1.04-1.14) | 1.58, (1.50-1.65) |
| Historical Average (4 year) | 295.4 | **59.8, (56.8-62.9)** | **0.92, (0.90-0.94)** | 1.28, (1.24-1.33) |
| Weighted Historical Average | 303.5 | **61.4, (56.1-66.9)** | **0.94, (0.91-0.98)** | 1.32, (1.27-1.38) |
| Poisson GLM | 304.7 | **61.6, (56.4-66.8)** | **0.95, (0.91-0.98)** | 1.32, (1.25-1.39) |
| Poisson GLM +SVI | 301.3 | **61.0, (57.4-65.1)** | 1.10, (1.04-1.16) | 1.38, (1.30-1.47) |
| Gradient Boosted Trees +SVI | 293.6 | **59.5, (55.9-63.1)** | **0.92, (0.89-0.96)** | **1.24, (1.18-1.30)** |
| GP | 287.2 | 58.2, (55.2-61.2) | **0.93, (0.91-0.95)** | **1.28, (1.23-1.34)** |
| CASTNet +SVI | 268.0 | 54.4, (52.0-56.6) | 1.07, (0.99-1.16) | 1.47, (1.36-1.58) |
| BST: Bayesian spatiotemporal | 301.0 | **61.1, (58.6-63.3)** | 0.95, (0.93- 0.96) | 1.26, (1.24-1.28) |
| BST + SVI | 305.4 | **62.0, (60.0-63.7)** | **0.92, (0.91-0.94)** | **1.23, (1.21-1.25)** |
| NBSpLag + SVI | 305.4 | **62.0, (60.3-64.0)** | **0.92 (0.89-0.95)** | 1.31, (1.27-1.35) |

Here BPR (higher is better) is our new intervention-aware metric, computed assuming an intervention budget for K=100 of 1620 possible tracts in Massachusetts. MAE (lower is better) and RMSE (lower is better) are common error-based metrics. For each metric, the best model mean is bolded, along with any other model whose uncertainty interval overlaps this mean (intervals are computed via resampling methods). The column titled “Total Overdoses Identified in top 100 tracts” contains the *true* number of observed fatal overdoses in the top 100 locations identified by the corresponding model.
Abbreviations: **BPR**: Best Possible Reach. **MAE**: Mean Absolute Error. **RMSE**: Root Mean Squared Error. **SVI**: Social Vulnerability Index covariates. **GLM**: Generalized Linear Model. **GP**: Gaussian Process. **NBSpLag**: Negative binomial regression with spatially-lagged features. **BST**: Bayesian spatiotemporal model

**Table 2.**
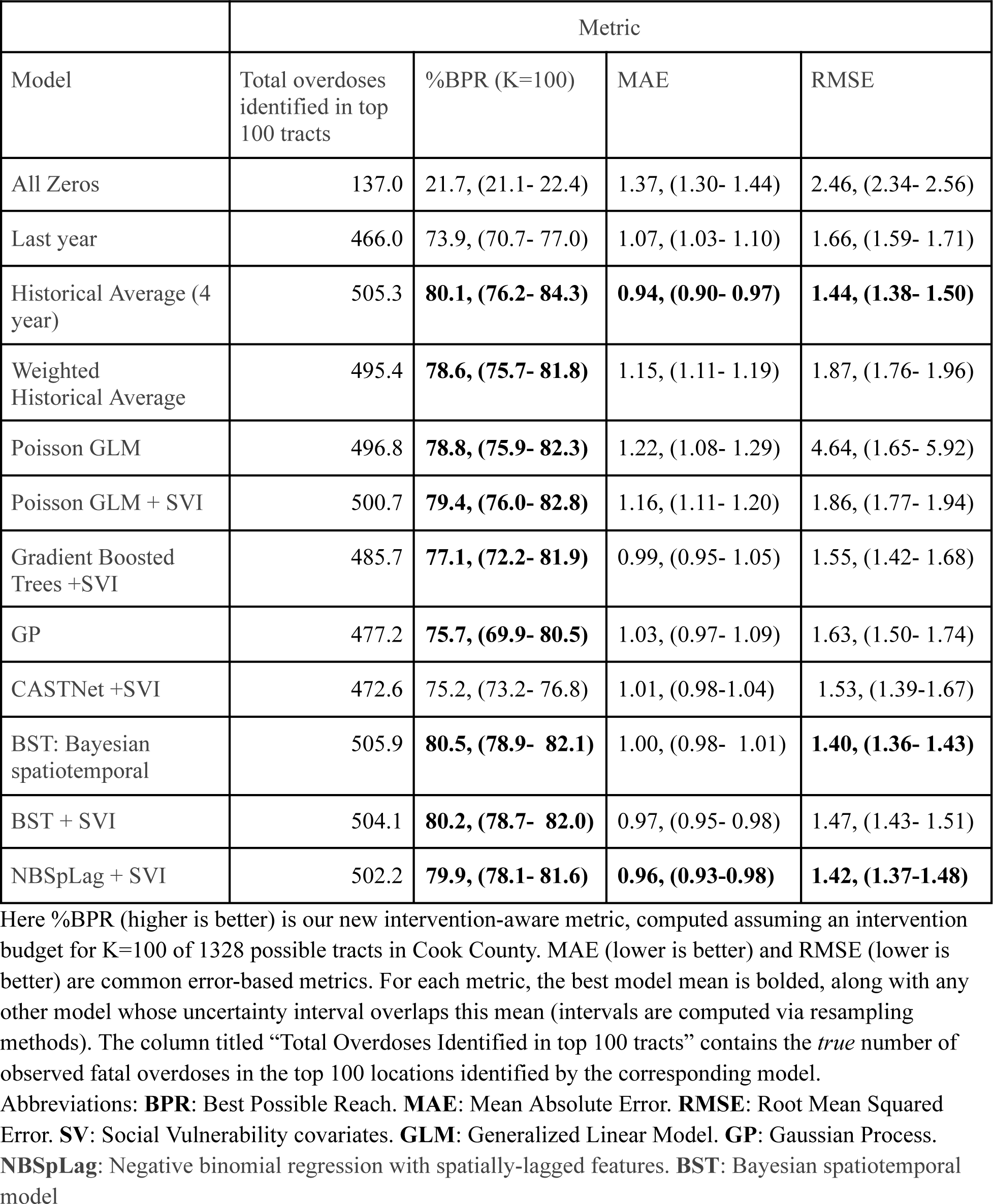
Comparison of fatal opioid-related overdose prediction models trained on Cook County, Illinois decedent data, from 2015 to 2020, then evaluated on data from 2021 and 2022.

In both catchment areas, we see reasons to prefer our proposed BPR metric to alternatives when the goal is effectively prioritizing where to intervene. First, in Massachusetts, both the Gaussian Process (GP) and Bayesian Spatiotemporal model (BST) have top performance as assessed by MAE and RMSE. However, the BST has higher %BPR than the GP (62.0% compared to 58.2%). Interventions guided by the BST model would have the potential to preemptively identify 18 additional fatal overdoses per year in the top 100 census tracts. Similarly, in Cook County the Gradient Boosted Trees model with SVI covariates has superior MAE and RMSE to the GLM model, yet has worse %BPR (77.1% versus 79.4% for GLM). Interventions guided by the GLM model (preferred via the %BPR metric) could reach 15 more fatal overdose events annually in Cook County.

We also observe that while complex models like BST do well in both catchment areas, so does the simple *historical average* baseline and its weighted extension. In Massachusetts, *historical average* delivers a BPR and MAE that fall within the uncertainty intervals of the best performing models. The weighted extension’s ultimate %BPR is so close to the top method (NPSpLag) that the difference amounts to identifying fewer than 2 additional overdose events annually. In Cook County, the *historical average* baseline delivers competitive scores (within the intervals of the best models) as assessed by all three metrics (BPR, MAE, and RMSE); the best model (BST) here would reach less than 1 additional overdose event annually.

## Discussion

Our study’s first contribution to the science of spatiotemporal forecasting of opioid-related overdose deaths is highlighting the need for extensive comparisons to a robust suite of simple baselines. This lesson matches reports^34,35^ from across the sciences, especially efforts in health^36,37^ and the social sciences^38^, that suggest advanced modeling techniques may not substantially outperform simpler baselines on some difficult prediction tasks. Our findings are similar in both the large state of MA and the far denser Cook County. In each catchment area, across both intervention-aware and error-based metrics, we found that a historical average baseline performed competitively (within the uncertainty bounds of top-ranked statistical models). The key to success here is careful selection of the number of recent years in the look-back period, following standard best practices for hyperparameter tuning.^39,40^ If this simple baseline model yields such high performance, it raises questions about the rationale for adopting more complex counterparts that require specialized expertise. Many prior overdose forecasting studies^20,23^ completely omit such baselines, or often include only the poor performing ones such as the last-year^28^ model or a too-long historical average^22^. For all future studies of opioid overdose forecasting, we recommend including historical averages with tuned look-back periods.

Our second contribution, developed in parallel to contemporary work^30^, is a new metric – percentage of best possible reach (%BPR) – which evaluates predictions based on their utility for informing decisions about where to intervene. In both Massachusetts and Cook County, we demonstrate that using %BPR as an evaluation metric can lead to different model rankings and different recommendations of where to intervene than error-based metrics like RMSE, improving the total number of annual fatal overdose events that could be preemptively identified by 15 in IL and 18 in MA. This is an important finding, as we believe that intervention-aware metrics like %BPR more closely reflect how public health agencies wish to use forecasting models to inform their intervention strategies.^20^

Lastly, we emphasize that our study is designed to be reproducible and open to extensions by other researchers. We released the software for fitting all models and computing all metrics under a permissive open-source license (link in Introduction). We also released our cleaned version of the public-domain Cook County dataset as well as all preprocessing code. Historically, overdose forecasting studies have not often shared code and have focused on private bespoke datasets, often reasonably due to privacy issues around decedent data. Enabling diverse researchers to pursue a common prediction task, especially via the availability of a public dataset for evaluation, has been a key driver of progress in predictive modeling^41^.

### Limitations

This study has several limitations. First, our findings come from only two places (Massachusetts and Cook County), and may not be generalizable to other counties, states, or public health jurisdictions. Cook County is predominantly urban, while Massachusetts is a large state with substantial urban, rural and suburban areas. The spatiotemporal trends in opioid-related mortality could thus be dramatically different in these two locations, necessitating different model rankings and intervention strategies. Furthermore, we acknowledge that not all Cook County deaths are reported to the Medical Examiner. The Medical Examiner’s jurisdiction only covers specific fatalities for cause-of-death determination.

Second, there are limitations to our analysis of the proposed BPR metric. For simplicity, all results here assumed an intervention budget of K=100 census tracts. Different K values may lead to different method rankings. Our suggested BPR metric is intended for identifying where to intervene to relieve high overall burden. However, it does not directly prioritize the rate of change. Interventions aimed to reduce risk in communities that are at very high risk but do not already have a high burden may not be correctly identified using BPR.

Finally, other choices of covariates are possible. Our focus on a limited set of covariates, derived from the SVI of the American Community Survey, was an intentional choice to ensure the nationwide availability of these covariates. Certain jurisdictions may possess useful alternate data sources, such as emergency medical service (EMS) calls, insurance claims data, and measures for a mix of linked administrative datasets^42^, necessitating additional covariate consideration for enhanced model performance.

## Conclusion

In an effort to better predict future fatal opioid-related overdose spikes and inform future harm-reducing interventions, we compared overdose forecasting options. Our study reinforces the value of intervention-aware metrics like %BPR in evaluating models for opioid overdose mortality forecasting. Our study also suggests that simple baselines like (weighted) historical averages should be included in future analyses, as more sophisticated and expensive-to-train models may not substantially outperform these baselines. As the opioid crisis continues to evolve, we hope our findings and our open-source resources enable improved model comparisons and better data-informed public health interventions that ultimately reduce the harm caused by overdose events.

## Data Availability

We share pre-processed data for the Cook County dataset, which is freely available. This data and all software is available at https://github.com/tufts-ml/opioid-overdose-models. The Massachusetts data belongs to the Massachusetts Department of Public Health

## Acknowledgments

We are grateful to both the Massachusetts Department of Public Health (DPH) and the Cook County Medical Examiner’s Office for data access.

Author KH was supported by the U.S. National Science Foundation under NSF award NRT-HDR 2021874. Author JK’s effort during a summer research program for undergraduates hosted at Tufts University was supported by NSF award REU-2149871. Authors KH, TJS, and MCH gratefully acknowledge support for early work on this project from NSF award IIS-1908617.

## Supplementary Material

### Additional Model details

Model hyperparameters are selected using the year prior to the test years as a validation year: 2019 for Massachusetts and 2020 for Cook County. When validating, models are trained through the year prior to the validation year. For evaluation on the test years, models are retrained through the validation year. Hyperparameters are selected by selecting the model with the highest BPR. In Massachusetts, we consider using up to 10 years of historical data when training (2010-2019). In Cook County there are fewer years of available historical data, and so training is limited to 2016-2020.

### All-Zeroes

The All-Zeroes model is presented to highlight two things: the BPR of a naive policy, and the RMSE and MAE of a naive model. This model is very simple: every prediction is always 0 fatal overdoses. However, this presents a challenge for calculating BPR: what are the top K locations if every location is tied? In this case, we take 10,000 samples, and randomly pick the K locations to serve as the numerator for BPR. We then calculate the BPR for each of 10,000 samples, and report the average.

### Last Year

For this model, the prediction is simply the previous year’s fatal overdose count. When predicting for the second evaluation year (2021 in Massachusetts and 2022 in Cook County), the fatal overdose count from the first evaluation year is used. This is subtly different from the behavior of the regression models, where the models are trained using no data from the evaluation years.

### Historical Average

In this model, the output is an un-weighted average of historical mortality. For both Massachusetts and Cook County 4 years are selected. These are both selected via the validation year. When predicting for the second evaluation year (2021 in Massachusetts and 2022 in Cook County), the fatal overdose count from the first evaluation year is used. This is subtly different from the behavior of the regression models, where the models are trained using no data from the evaluation years.

### Weighted Historical Average

This model is a linear regression on historical fatal overdose count using only past years mortality as a predictor. Scikit-learn’s^43^ ridge regression is used, which performs L2-regularized regression. The regularization strength α is selected via hyperparameter search by trying 29 evenly spaced values on a log scale between 10^-6^ and 10^8^. For Massachusetts, 10 years of historical data and an α of 10^4.5^ is used. For Cook County, 3 years of historical data are used and an α of 10^4.5^ is selected.

### Linear Poisson GLM

This uses Scikit-learn’s^43^ Generalized Linear Model with a Poisson Likelihood and a log link. The hyperparameters explored are the number of prior years of mortality to include in the model and the L2 regularization strength α. Up to 10 years of previous mortality were considered for Massachusetts and 6 years for Cook County. The model is run with and without social vulnerability covariates. **Without social vulnerability covariates** In Massachusetts 10 years of prior mortality are used in the model with an α of 1. In Cook County, 5 years of historical data are used with an α of 10^0.5^. **With social vulnerability covariates** In Massachusetts 6 years of prior mortality are used in the model with an α of 1. In Cook County, 5 years of historical data are used with an α of 10.

### Gradient Boosted Trees

This uses Scikit-learn’s^43^ Histogram-basedGradient Boosted Trees model. We considered both squared-error and Poisson loss functions. We tested using both 32 and 128 maximum iterations. For the minimum samples per leaf we used 9 equally spaced values between 2^0^ and 2^8^ on a log-2 scale. The maximum number of leaf nodes tested were 5 equally spaced values between 2^4^ and 2^8^ on a log-2 scale. Up to 10 years of previous mortality were considered for Massachusetts and 6 years for Cook County.

### Gaussian Process Models

Here we use Scikit-learn’s^43^ Gaussian Process (GP) implementation. Due to the high computational cost of GP models, and following prior work^20^, we only consider up to 5 years of historical data for both Massachusetts and Cook County, and we omit social vulnerability covariates. As in the prior work,we use a kernel that additively combines a Radial-Basis Function (RBF) kernel with a white noise kernel. The initial length scale of the RBF kernel is set to 0.5, and the noise level bounds on the white noise are set to (10^-5^, 10^1^). The outcomes are normalized to 0-mean and unit variance. Up to 9 restarts of the optimizer are used.

### CASTNet

While we attempted to follow the original implementation of CASTNet^22^ as closely as possible, there are some significant modifications. The original CASTNet was concerned with predictions at a fine temporal-resolution, weekly. However, our initial experiments found little benefit at this scale, and we consider a much coarser scale: annual predictions. We lack the high-resolution crime data that the original CASTNet project uses as dynamic covariates. Furthermore, while we do have demographic and economic variables (the 5-dimensional Social Vulnerability index), these are not static at the annual scale but dynamic, accordingly these are used as the only dynamic covariates. For static covariates, only the latitude and longitude of the census tract are used. We use the hyperparameters selected by the original work: the LSTMs have a hidden unit size of 32 with a dropout value of 0.1, the group-level regularization coefficient is 0.0025, and the optimizer used was Adam with a learning rate of 0.5. Given that we have 2 evaluation years, we train the model twice, once with a lag time of 1-year and again with a lag time of 2-years. The 1-year lag model is used to predict for the first evaluation year (2020 in MA and 2021 in Cook County) and the 2-year lag model is used to predict for the second. This way no training data leaks into the model.

### Bayesian Spatiotemporal Models

In the original work^23^ on Bayesian Spatiotemporal models for opioid overdose forecasting, three separate models are proposed. All three models use an Autoregressive-1 term to model temporal dependence, and two of the models use spatial correlations. Because all three models are reported to behave similarly, we use what is called “Model 1”, lacking spatial correlations. Furthermore, the authors state that any number of temporal terms could be used, but do not specify which. Here, we choose a linear temporal term.This model is implemented using R-INLA^44^. Linear coefficients are used when adding the social vulnerability covariates.

### Negative Binomial Regression with Spatially Lagged Covariates

The authors of this method^28^ helpfully provide code to run this model which we were able to use with little modification. Census tract level population estimates are taken from the same survey data as the social vulnerability covariates. The carrying capacity is initialized to 5% of the population in the first year of training data (2010 for MA, 2015 for Cook County).

## Notes

### Competing Interest Statement

The authors have declared no competing interest.

### Funding Statement

Author KH was supported by the U.S. National Science Foundation under NSF award NRT-HDR 2021874. Author JKs effort during a summer research program for undergraduates hosted at Tufts University was supported by NSF award REU-2149871. Authors KH, TJS, and MCH gratefully acknowledge support for early work on this project from NSF award IIS-1908617.

